# Gene-Pseudogene Inversions as a Hidden Source of Missing Heritability

**DOI:** 10.1101/2025.10.01.25336578

**Authors:** Ilaria Quartesan, Stefano Facchini, Arianna Manini, Ricardo Parolin Schnekenberg, Chiara Pisciotta, Stefania Magri, Sara Negri, Carlo Gaetano, Adriana Rebelo, Jacquelyn Schatzman Raposo, Radim Mazanec, Riccardo Curro, Natalia Dominik, Stephanie Efthymiou, Matilde Laurà, Tiffany Grider, Shawna ME Feely, Vera Fridman, Alessandro Bertini, Gustavo Maximiano Alves, Lucia Ferullo, Arianna Ghia, Claudio Caccia, Francesca Balistreri, Paola Saveri, Luca Crivellari, Isabella Moroni, Federica Rachele Danti, Tiziana Mongini, Franco Taroni, Michaela Auer-Grumbach, Enrico Bugiardini, James N Sleigh, Arianna Tucci, Henry Houlden, Petra Laššuthová, Pavel Seeman, Anna Basile, Elisa Giorgio, Michael E Shy, Stephan Zuchner, Mary M Reilly, Davide Pareyson, Andrea Cortese

## Abstract

Historically defined as non-functional copies of coding genes, pseudogenes are an abundant yet underexplored element in the human genome, despite growing evidence linking them to human diseases. From a genome-wide screen, we identified 411 gene-pseudogene pairs located in opposite orientation, an arrangement which is permissive for the occurrence of inversions, including 46 genes already associated with human disease. Next, by analysing long read sequencing (LRS) data from the 1000 Genomes Project, we confirmed that at least 3.6% of healthy individuals carry an inversion involving one of these gene/pseudogene pairs, while they were previously undetected by short read sequencing.

Most importantly, we identified novel and recurrent inversions between *SORD* and its pseudogene *SORD2P* in 13 out of 151 patients (9%) affected by *SORD*-related Charcot–Marie–Tooth (CMT) neuropathy, including 6 out of 8 (75%) of *SORD*-CMT cases where only one pathogenic variant was identified on short read sequencing, making it the third most common pathogenic allele causing *SORD*-CMT. Of interest, gene-pseudogene pairs displaying chromatin contact in Micro-C data, including *SORD/SORD2P*, were found to be more likely to undergo inversion events. Overall, our results highlight gene–pseudogene inversions as a previously underrecognized type of pathogenic structural variant. Wider use of LRS could reveal their true prevalence and contribution to the missing heritability in Mendelian diseases.

## Main

Pseudogenes are repetitive DNA sequences that originate from the duplication or retrotransposition of protein-coding genes but lose their protein-coding ability due to the progressive accumulation of deleterious mutations under neutral selection. Once thought to be non-functional genomic relics, they are now increasingly recognized for their regulatory functions and involvement in human diseases [1-2].

In particular, unprocessed pseudogenes maintain an exon–intron structure and a high sequence similarity to their parental genes, thus promoting recombination during meiosis. Short DNA segments can transfer from pseudogenes to their functional counterparts via gene conversion [3], a process that has been extensively studied and implicated in several hereditary diseases, including congenital adrenal hyperplasia [4], polycystic kidney disease type 1 [5], Gaucher disease [6], and spinal muscular atrophy (though *SMN2* is a hypomorphic *SMN1* paralog, not a true pseudogene) [7]. Unequal crossing-over can also lead to deletions or duplications of the parental gene, as seen in *BRCA1* deletions linked to breast and ovarian cancer susceptibility [8], or in *SUZ12/SUZ12P1* recombination events leading to *NF1* deletion in neurofibromatosis [9].

Conversely, inversions resulting from gene–pseudogene recombination seem rare, with the only documented examples being the *IDS/IDS2* inversion causing Hunter syndrome [10] and the inversions between intron 22 of *F8* and one of its two homologues (*int22h2/int22h3*), which account for nearly half of all severe Haemophilia A cases [11]. However, given that short-read sequencing technologies are inherently limited in their detection of dosage-neutral structural variants, particularly when these occur in repetitive regions [12-13], such events may be systematically overlooked, and therefore more prevalent in the human genome than currently recognized.

### Identification of gene-pseudogene candidate pairs in inversion-permissive orientation

Prompted by this hypothesis, we first conducted a genome-wide analysis to identify gene– pseudogene pairs located on the same chromosome and arranged in an opposite orientation – a configuration permissive for the occurrence of inversion events mediated by non-allelic homologous recombination (NAHR) (***Figure 1a***) [14]. To do this, we analysed the *Psicube* dataset, which is a curated database of 10,371 known human pseudogenes originating from 3,707 unique parent genes [15]. We identified 411 pseudogenes (4% of all known pseudogenes) that stemmed from 270 parent genes (22.2% of parent genes have more than one pseudogene) – which met the described orientation requirement (***Supplementary Table S1***).

**Figure 1:**
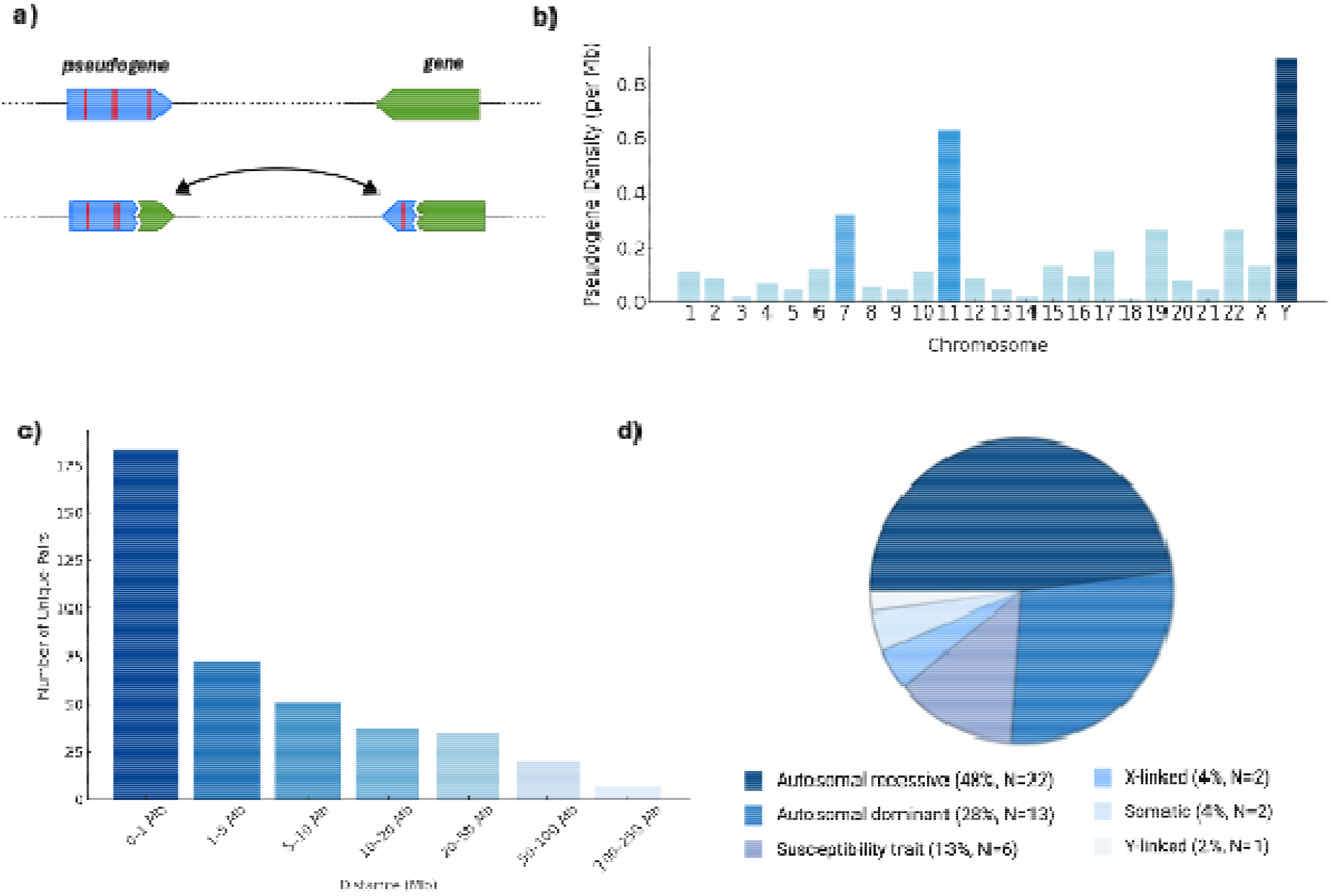
Genome-wide characterization of gene–pseudogene pairs located in inversion-permissive orientation. (a) Schematic illustration of the proposed inversion mechanism. The functional gene (green) and its pseudogene (blue, carrying deleterious mutations indicated by red bars) are positioned in opposite orientation (head-to-head/tail-to-tail). Pairing between the oppositely oriented duplicated sequences of the gene and pseudogene via non-allelic homologous recombination results in the inversion of the intervening genomic segment, potentially generating chimeric gene–pseudogene fusion sequences at the breakpoints. (b) Bar chart illustrating chromosomal distribution of gene–pseudogene density, calculated as the number of pseudogenes per base pair for each chromosome (c) Bar chart illustrating the distribution of genomic distances between pseudogenes and their parent gene, with each bin being left-inclusive and right-exclusive. (d) Cross-referencing with the OMIM database revealed that 46 parent genes are associated with known human diseases, among which autosomal recessive disorders represented the largest category.

Analysis of their genomic distribution showed that the identified pseudogenes are unevenly distributed across the genome (***Figure 1b***). Chromosome Y exhibited the highest density, likely attributable to its elevated mutation rate, absence of recombination mechanisms for repair, and reduced evolutionary constraint [16]. Chromosomes 7 and 11 also displayed a high concentration of duplicated pseudogenes due to the presence of large segmental duplications, particularly involving olfactory receptor genes [17-18]. Because NAHR is more likely to occur when homologous sequences are in proximity, we examined the linear distance between each pseudogene and its parental gene and found that 42% of identified pseudogenes lie within 1 Mb of their parent genes (***Figure 1c***). Gene Ontology (GO) enrichment analysis of the parent genes revealed a strong overrepresentation of genes involved in sensory perception and metabolic processes.

Notably, 46 (17%) of these genes are already associated with human diseases according to the OMIM database [19] (***Supplementary Table S2* and *Figure 1d***). This set includes genes, such as *PKD1* (polycystic kidney disease) and *PMS2* (Lynch syndrome), where gene conversion events involving their corresponding pseudogenes are established pathogenic mechanisms [5,20].

### Long-read sequencing data from the 1000 Genome Project reveals an increased frequency of gene-pseudogene inversions in healthy individuals

After identifying gene–pseudogene pairs arranged in an inversion-permissive orientation, we next sought to assess the actual occurrence of such inversions in a reference population, estimating both their frequency and genomic distribution. To this end, we performed a targeted analysis on the recently released resource of Oxford Nanopore Technology (ONT) whole-genome long-read sequencing (LRS) data of 1,019 individuals from the 1000 Genomes Project (1kGP)[21].Although large inversions (>50–100 kb) exceed typical read lengths, their presence can be inferred from split-read alignments that map discontinuously and in reversed orientation across breakpoints.

Leveraging a bioinformatics pipeline based on VacMap, a long-read alignment tool optimized for structural variant detection [22], and custom breakpoint filtering (see *Methods*), we initially identified 206 putative inversions involving 11 gene–pseudogene pairs (*Supplementary Table S3*). After rigorous manual curation (filtering out potential false positives and evaluating breakpoint consistency), we confirmed the presence of 37 high-confidence inversions spanning 9 distinct gene– pseudogene pairs (*Table 1*). All but one pair were located within 1 Mb of each other, with the size of inversions ranging from ∼56 kb to ∼5 Mb, which are significantly larger than typical LRS read lengths. Our results indicate that at least 3.6% of individuals carry one or more gene–pseudogene inversions, though this is likely an underestimate given the high stringency of our filtering criteria.

**Table 1:**
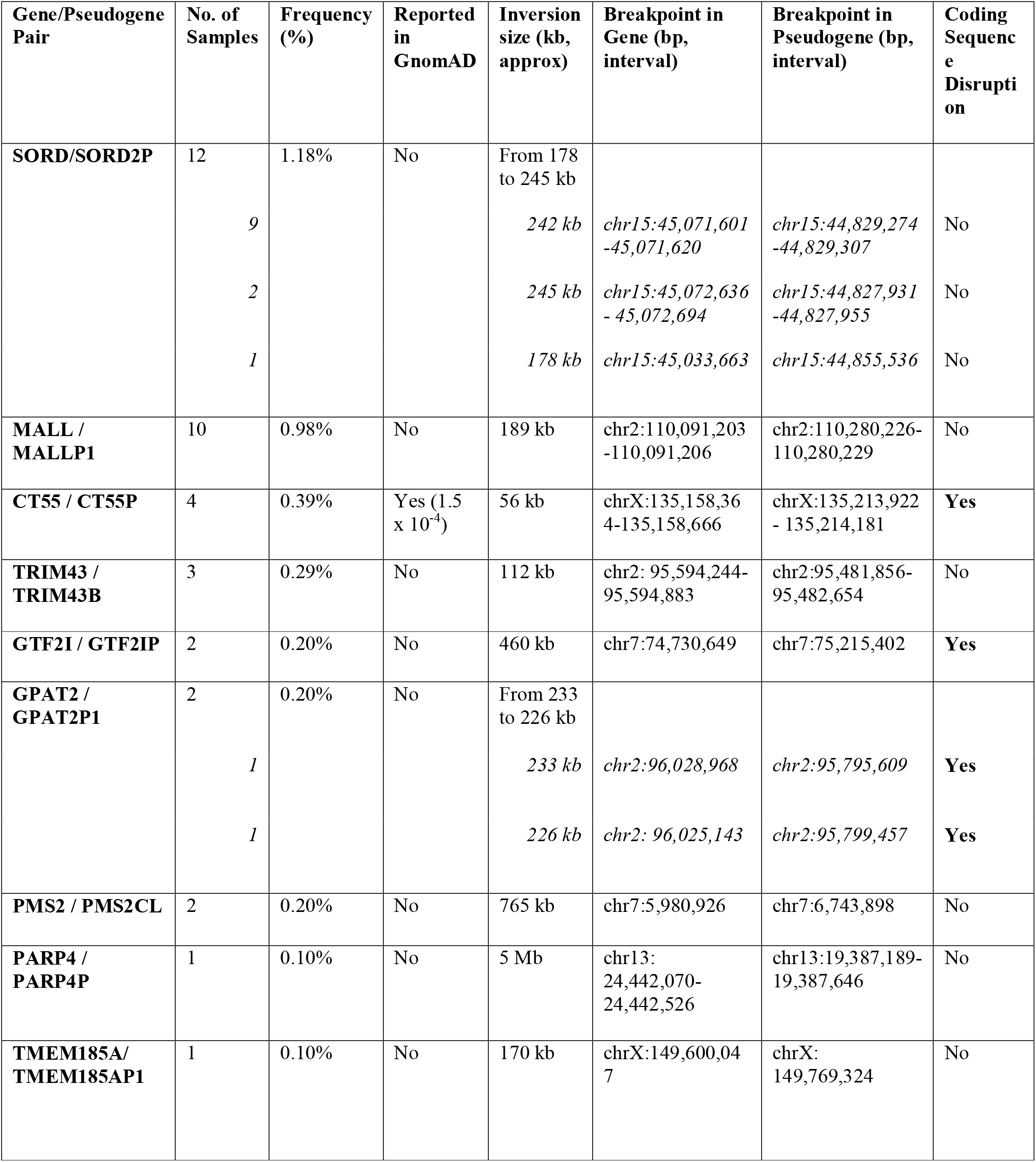
Long-read sequencing identifies recurrent gene–pseudogene inversions from the 1000 Genomes Project. The table summarizes gene–pseudogene inversion events identified through whole-genome long-read sequencing (LRS). Inversion events were clustered based on breakpoint proximity, using a ±1 kb tolerance to define distinct events. For each inversion, the number of carriers, approximate size, and breakpoint (reported as ranges to reflect the inter-sample variability) in both the gene and pseudogene are shown. Coding impact refers to predicted disruption of the canonical transcript by frameshift or exon loss.

Importantly, only one of the validated inversion events (*CT55*/*CT55P*) is reported in the gnomAD structural variant (SV) database, with a minor allele frequency (MAF) of 1.5 × 10□□, thus substantially lower than our observed frequency. This discrepancy underscores the known limitations of short-read sequencing in detecting inversions, particularly within regions of high sequence homology. While most of the identified inversions appeared to be neutral, eight of them in three genes, *CT55, GTF2I* and *GPAT2*, were predicted to alter the coding sequence of the parent gene through frameshift or exon deletions.

Several inversions involved clinically relevant genes, including *PMS2* [20] and *GTF2I*, a gene located within the 7q11.23 region that is deleted in Williams syndrome and of functional relevance to the disease’s cognitive and social phenotype [23]. Notably, the *SORD*/*SORD2P* locus exhibited the highest number of recurrent inversion events (12 alleles). Although none of the *SORD*/*SORD2P* inversions identified in the 1kGP dataset were predicted to disrupt the *SORD* reading frame, this finding is particularly noteworthy, as biallelic loss-of-function variants in *SORD* are a common cause of inherited neuropathy [24]. This led us to speculate that additional inversions leading to SORD frameshift may exist and represent the underlying cause of progressive nerve degeneration in genetically unsolved individuals affected by Charcot–Marie–Tooth disease (CMT) or distal hereditary motor neuropathy (dHMN).

### Inversions between *SORD* and *SORD2P* are a common cause of inherited neuropathy

To explore this possibility, we investigated three individuals with a clinical diagnosis of CMT and elevated fasting serum sorbitol levels—a reliable biomarker of an alteration in the polyol pathway caused by loss of *SORD* enzyme function [24]. In each case, short-read whole-genome sequencing (srWGS) identified only a single heterozygous pathogenic variant in *SORD* (c.757delG, p.Ala253Glnfs*27), suggesting the presence of a second, undetected pathogenic allele.

We performed LRS on all three cases, but neither single nucleotide variant (SNV) analysis or structural variant callers revealed any promising candidates. However, by leveraging the same targeted split-read-based approach and careful visual inspection adopted in our genome-wide study, we uncovered a recurrent inversion between *SORD* and its pseudogene *SORD2P* in all three individuals. (***Figure 2a***) Although precise breakpoint mapping was hindered by the high homology of the region, we determined that the inversion spans approximately from chr15:44,844,906 (*SORD2P*, intron 5) to chr15:45,067,711 (*SORD*, intron 4). The inversion therefore replaces the first 4 exons of *SORD* with the first 5 exons belonging to *SORD2P*, which harbour multiple nonsense variants. Retrospective analysis of srWGS data confirmed that this *SORD/SORD2P* pathogenic inversion, like most inversions identified in this study, was completely undetectable through short-read technologies (***Figure 2a***).

**Figure 2.**
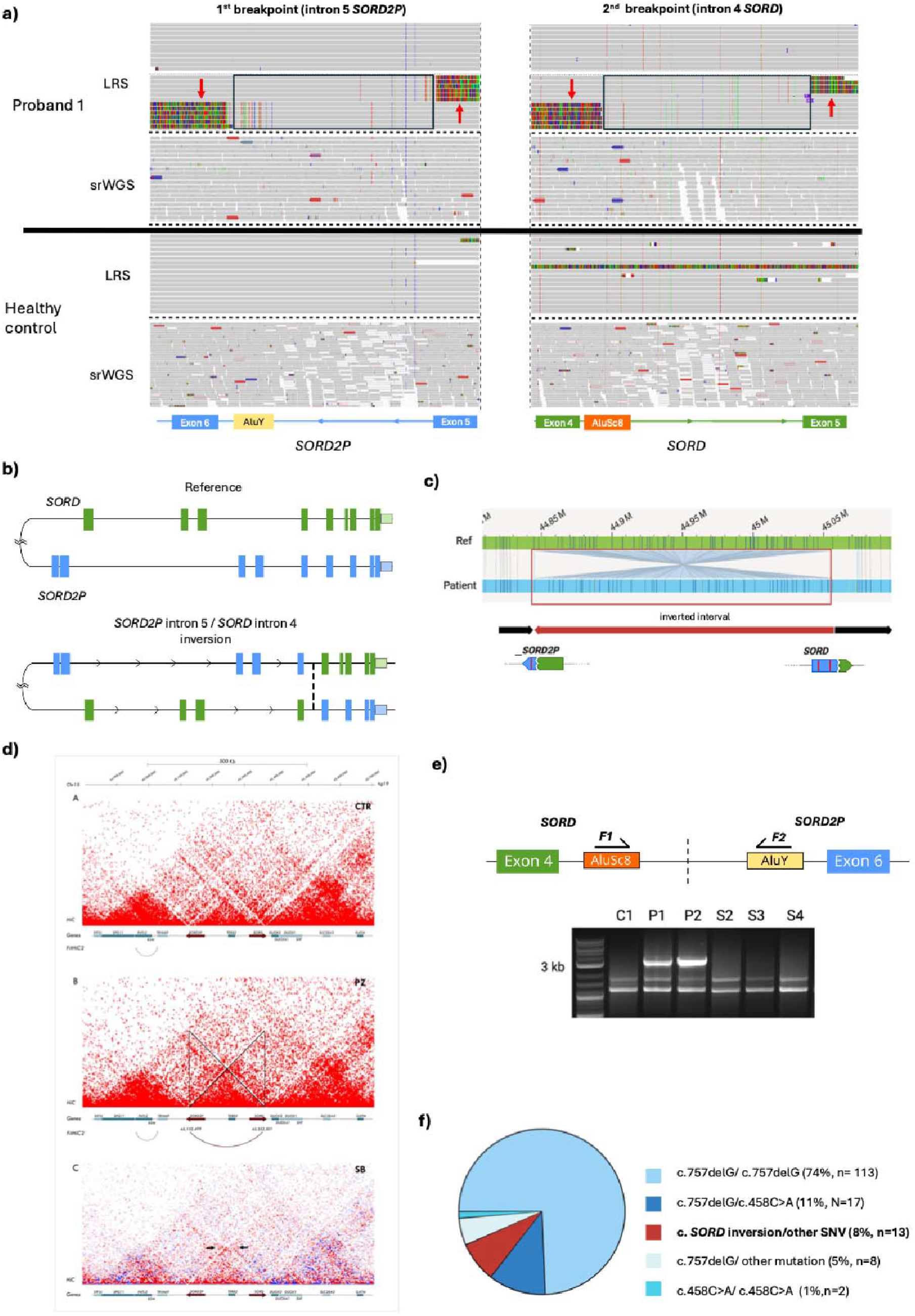
Identification of SORD/SORD2P inversion as a common pathogenic allele in CMT-SORD. a) IGV visualization of the inversion breakpoint (intron 5 SORD2P and intron 4 SORD). Split and soft-clipped long-reads (red arrows) identify insertions/deletions of Alu elements specific to SORD or SORD2P, thus delineating the genomic interval of the breakpoints of the inversion between SORD and SORD2P (black boxes). Short-read sequencing fails to resolve the inversion due to high sequence homology. b) Schematic representation of the inversion’s genomic effect: the first four exons of SORD are replaced by the first five exons of SORD2P, which harbor multiple nonsense variants, leading to non-sense mediated decay and loss of SORD function. c) Optical Genome Mapping (OGM) of the inversion. The first breakpoint is located within the SORD2P pseudogene, while the second is within SORD. d) Zoom-in of the Hi-C ∼1 Mb region surrounding the SORD-SORD2P locus (skin fibroblasts; hg19; 5 kb resolution; raw counts) in a control (panel A; CTR) and the patient (panel B; proband N, SORD-CM). Statistically significant chromatin contacts identified by FitHiC2 are shown for both the control and patient samples. A patient-specific interaction (dark red curve, panel B) between chromosome 15 positions 45,132,499 and 45,352,501 was detected, consistent with the SORD/SORD2P inversion identified in the patient. Panel C displays a heatmap obtained with the subtraction method (SB map; patient - control), in which the inversion is visualized as a characteristic bow-tie pattern located distant from the matrix (arrows). e) Schematic representation of the breakpoint-specific PCR assay (top) and corresponding gel electrophoresis of PCR products from SORD-CMT patients and controls. Primers target divergent Alu elements in SORD2P intron 6 and SORD intron 5. In the reference genome, primers F1 and F2 are oriented in the same direction and ∼226 kb apart, precluding amplification. Upon inversion, F1 now lies ∼3.2 kb from F2 and in opposite direction, thus enabling amplification of a PCR product. Gel electrophoresis of breakpoint PCR assay shows the presence of a 3.2 Kb PCR product in probands P1 and P2 (heterozygous for SORD c.757delG and the inversion), but not in control (C1) or in probands that do not carry the inversion (S1–S3). f) Pie chart depicting the prevalence of SORD variants in a cohort of 156 CMT-SORD patients. SORD/SORD2P inversions represent the third most common pathogenic allele.

Given that the size of the inversion (∼200 kb) far exceeded the span of single long-read molecules, we complemented our analysis with optical genome mapping (OGM) to validate its presence. OGM provided unambiguous, direct visualization of the *SORD/SORD2P* inversion in all three cases (***Figure 2b***). This inversion was absent from the Bionano control database of 362 individuals and from our in-house dataset of 189 individuals.

Finally, we generated Hi-C data from one affected individual, which also confirmed the presence of the inversion as an increased *SORD/SORD2P* interaction, (***Figure 2c***), and thus highlighting the possible diagnostic utility of Hi-C for the detection of this peculiar type of SV.

Given the presence of a recurrent breakpoint region limited by distinct Alu elements that are part of SORD or SORD2P genes, we developed a breakpoint-specific PCR assay to further investigate the frequency of the *SORD/SORD2P* inversion in a larger cohort of CMT patients carrying only one known pathogenic *SORD* variant. Primers were designed to target the unique Alu elements within *SORD2P intron 6* and *SORD intron 5*, and to generate a product only if the inversion brings these retrotransposons into proximity and correct orientation (***Figure 2d)***. This method confirmed the presence of the inversion in all three cases, while showing its absence in controls.

We subsequently screened, using the same PCR-based method, 38 additional individuals with CMT/dHMN who carried a single known pathogenic *SORD* variant, including 8 for whom serum sorbitol levels were available and elevated, and identified 9 new carriers of the *SORD/SORD2P* inversion. Notably, among these 8 individuals with elevated sorbitol, 5 (62.5%) carried the inversion. We next performed LRS in the three patients with elevated serum sorbitol and only one pathogenic *SORD* variant on srNGS, who nonetheless tested negative for the recurrent inversion. Strikingly, in one case we identified a second, distinct *SORD/SORD2P* pathogenic inversion (breakpoints: *SORD intron 3* at chr15:45,044,084; *SORD2P intron 4* at chr15:44,846,734 that, like the more common *SORD* intron 4 / *SORD2P* intron 5 inversion, disrupts the *SORD* coding sequence, providing further evidence of the locus’s susceptibility to inversion events.” ***(Supplementary Figure 1)***. In the two remaining cases, LRS unveiled two pathogenic SNVs — c.142G>T (p.Asp48Tyr) and c.1021G>T (p.Gly341Ter) —which were missed by srNGS likely due to ambiguous mapping of reads between highly homologous regions.

The phenotype of individuals carrying the *SORD/SORD2P* inversion was consistent with that observed in *SORD*-related CMT caused by biallelic point mutations [24]. Disease onset typically occurred during the second decade of life and was characterized by distal lower limb weakness with minimal sensory involvement. Nerve conduction studies (NCS) confirmed a pure or predominantly motor neuropathy in all cases, in line with the clinical phenotype. Detailed clinical, demographic, and electrophysiological data are available in ***Supplementary Table 4***.

Importantly, the identification of pathogenic *SORD/SORD2P* inversions had direct clinical implications, enabling the inclusion of two affected individuals in the ongoing clinical trial evaluating the safety and efficacy of govorestat, a novel aldose reductase inhibitor [25].

Remarkably, *SORD/SORD2P* inversions represent the third most common pathogenic *SORD* allele, after c.757delG (p.Ala253Glnfs*27) and c.458C>A (p.Ala153Asp) (***Figure 2e***) [26]. This finding highlights gene–pseudogene inversions as a potentially frequent and underrecognized cause of human disease, systematically missed by short-read technologies but reliably detected through LRS and OGM.

### Higher-order chromatin structure may promote gene–pseudogene recombination events

The recurrent nature and the prevalence of *SORD/SORD2P* inversions prompted us to explore potential factors predisposing this locus to structural rearrangements. It has been previously speculated that higher-order genome organization contributes to the occurrence of chromosomal rearrangements and SVs by bringing susceptible breakpoint regions into close spatial proximity, thereby increasing the likelihood of interactions [27-28].

To assess the contribution of local chromatin architecture at the *SORD/SORD2P* locus, we analysed publicly available Hi-C and Micro-C datasets [29-30]. While Hi-C did not reveal any notable structural features, Micro-C data showed a distinct “stripe” of elevated contact frequency extending symmetrically across *SORD, SORD2P*, and their intergenic region (*Figure 3a*). This pattern is consistent with the formation of a chromatin loop that brings these homologous loci into close spatial proximity (*Figure 3b*), potentially predisposing them to inversion through NAHR.

**Figure 3.**
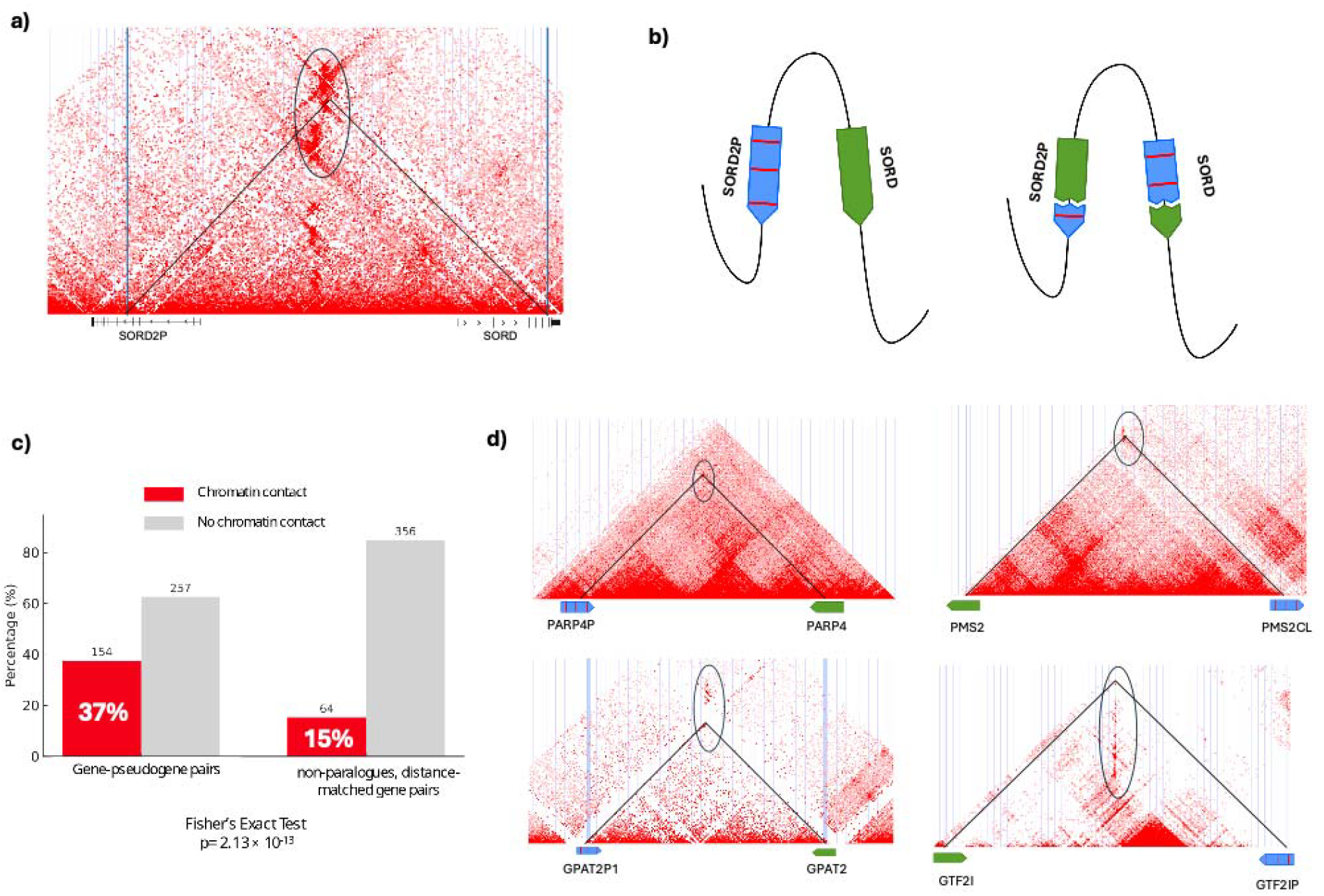
Increased 3D chromatin contacts between gene/pseudogenes undergoing inversion or in inversion-permissive orientation. (a) High□resolution chromatin interaction (Micro□C) matrix illustrating the genomic region encompassing SORD2P and SORD. Vertical bars/lines correspond to the location of the breakpoint of the inversions. The circled area highlights the continuous region of high contact frequency; the intersection point of the two breakpoint coordinates falls within this domain. (b) Schematic representation of the possible looping that brings SORD and SORD2P into physical proximity, which may facilitate recombination events between the two loci. (c) Bar chart comparing the proportion and absolute number of gene–pseudogene pairs (left) versus non-paralogues, distance-matched gene pairs (right) that exhibit significant chromatin contact after Bonferroni correction (p < 1e-6). Each group is divided into two bars: pairs in significant contact (red) and pairs not in contact (grey). The enrichment of significant contacts among gene–pseudogene pairs compared to controls is statistically significant (Fisher’s Exact Test p < 1 × 10□^12^). (d) Micro-C contact matrices for four additional gene/pseudogene pairs where inversions were identified. The circled regions indicate the domains of high contact frequency encompassing the inversion breakpoints.

To determine whether this observation was specific to *SORD* or reflective of a broader pattern among gene–pseudogene pairs, we leveraged FitHiC2, a computational tool for detecting statistically significant chromatin interactions, [31] to analyze our list of 411 gene–pseudogene pairs. We found that 37% (154/411) of these pairs displayed significant chromatin contact, compared to only 15% (64/420) of randomly selected, distance-matched, and non-paralogues gene pairs (Fisher’s Exact Test, p < 1 × 10□^12^) *(Figure 1c)*. This enrichment suggests that chromatin interactions between gene–pseudogene pairs are non-random, likely facilitated by the sequence homology, and may contribute to the recurrence of structural rearrangements. Notably, among the eight gene–pseudogene pairs, other than *SORD/SORD2P*, in which inversions were identified by LRS in the 1000 Genome Project control database, six exhibited significant chromatin contact based on FitHiC2-derived interaction scores. Visual inspection of the Micro-C tracks further revealed that, in four of these cases, a “stripe” of elevated chromatin contact spanned the breakpoint coordinates, resembling the pattern observed at the *SORD/SORD2P* locus (*Figure 1d*).

### Conclusions

Our findings establish gene–pseudogene inversions as an underrecognized yet clinically relevant class of SVs that are mostly invisible to short-read technologies. We identified hundreds of inversion-permissive gene–pseudogene pairs and showed that a subset undergoes recurrent rearrangements in the general population. Notably, we also demonstrated that inversions involving *SORD* and its pseudogene *SORD2P*, including a recurrent inversion between *intron 5* of *SORD2P* and *intron 4* of SORD, represent the third most common pathogenic alleles in CMT-SORD, highlighting the importance of screening for the inversion in patients presenting with elevated sorbitol levels, but lacking a “second hit”, either by PCR or LRS. More broadly, similar pathogenic inversions may exist in *SORD/SORD2P* as well as at other clinically relevant loci but remain largely undetected due to the technical shortcomings of srWGS.

Some caveats should be noted in relation to this study. While LRS improves structural variant detection in repetitive regions compared to srWGS, it remains error-prone, requiring stringent filtering and manual curation. Additionally, while the common *SORD/SORD2P* inversion was confirmed via OGM and breakpoint-specific PCR, similar validation was not possible for inversions detected as part of the 1000 Genomes Project, as we lacked access to DNA samples. Finally, although Micro-C is designed to exclude or downweight ambiguously mapped reads, a potential confounding effect of homologous regions on the detected contact signals cannot be entirely ruled out.

Despite these limitations, our findings highlight gene–pseudogene inversions as an underrecognized class of potentially pathogenic SVs and suggest that their systematic identification through novel sequencing technologies may help to resolve a portion of the missing heritability in Mendelian diseases.

## Methods

### Candidate inversion-permissive gene-pseudogenes identification from the psiCube database

We leveraged psiCube, an updated pseudogene resource that provides pseudogene annotation and information on the parent gene [15]. From this resource, we initially extracted a comprehensive set of 10,371 gene–pseudogene pairs. We then considered only pairs satisfying the conditions:

1. the pseudogene is unprocessed (specifically, we included only the following types defined in the catalog:”unitary_pseudogene”, “unprocessed_pseudogene”, “transcribed_unprocessed_pseudogene”, “pseudogene”)
2. Gene and pseudogene lie on the same chromosome and have opposite orientation.

### Analysis of long-read data from the 1000 Genomes Project

We leveraged the resource of 1,019 Oxford Nanopore Technologies (ONT) long-read genomes sampled within 26 human populations from the 1000 Genomes Project. [21]. The original publication and documentation contain detailed information on sample collection, sequencing protocols, and quality-control measures. The genomes were first realigned with VACmap, a novel long-read aligner specifically designed for complex structural variation discovery [22]. To improve processing time, only regions enclosing the genes and pseudogenes in the list (extended by 50 kbp on both flanks) were extracted from the original CRAM files and realigned, using the high error rate mode suitable for ONT sequencing (-mode H). Sniffles v2 [32] with standard parameters was then used to generate VCF files with SVs. From the VCFs and combining standard bcftools commands and a custom python filtering, we obtained a list of inversions where one breakpoint is in the gene and the other breakpoint in one of the corresponding pseudogenes, for a total of 2758 inversions in all samples. We then proceeded to apply a further filtering step, keeping only variants supported by at least 5 reads satisfying the following conditions: 1) the read is clipped; 2) the clipping position falls within 500 bp of the predicted breakpoint in the parent gene; 3) there is exactly one supplementary alignment involving the clipped sequence; 4) the supplementary alignment has one end within 500 bp of the predicted breakpoint in the pseudogene; 5) the primary and the supplementary alignment have opposite orientations. This filtering step was implemented via a python script using pysam v0.22.0. In this way, a final short list of 206 inversions in 180 samples was finally obtained. The variants were subsequently assessed by visual inspection in Integrative Genomic Viewer (IGV), which led to removal of low confidence inversions, including *CES1* (n=108) and *ARHGEF5* (n=55), where the mapping of the split reads to the pseudogene was ambiguous, because of the presence of an alternative haplotype, frequent in the general population, for the region of interest (chr16_GL383556v1_alt and chr7_KZ208913v1_alt, respectively for *CES1* and *ARHGEF5*). In both cases, the alternative sequence shows an even higher degree of similarity of the gene to the respective pseudogene, leading to an inaccurate read alignment.

### Enrolment of patients affected with inherited neuropathy and missing second variant in SORD after short-read analysis

Patients diagnosed with CMT2 or dHMN and carrying only one pathogenic variant in *SORD* were identified from the Queen Square Institute of Neurology (London, UK), the Fondazione IRCCS Istituto Neurologico “Carlo Besta” (Milan, Italy), and University Hospital Motol (Prague, Czech Republic), as well as patients within the 100,000 Genomes Project (100kGP) and the GENESIS platform. In biochemical tests, eight of the patients showed a raised serum sorbitol level, which is a highly specific biomarker of CMT-SORD. Informed consent was obtained from all participating subjects.

### Long-Read Sequencing (LRS)

DNA was extracted from blood using the Qiagen DNeasy Blood & Tissue kit, as per the manufacturer’s instructions. Quality control was performed using a Femto Pulse System (Agilent Technologies) and a Qubit fluorometer (Invitrogen). Sample preparation for Nanopore sequencing was performed using the ligation sequencing kit SQK-LSK110. DNA was sequenced on a PromethION using R9.4.1 flowcells (ONT). Samples for Probands 1-3 were run for 72□h including a washing and reload step after 24□h and 48□h. Library preparation and sequencing were performed by the UCL LRS facility. Reads in FASTQ format from both ‘fastq_pass’ and ‘fastq_fail’ folders were initially aligned to the hg38 reference with minimap2, and processed with DeepVariant and Sniffles2, for SNV and SV detection, respectively. For Proband 1, the region encompassing SORD and SORD2P was re-aligned with ‘bwa mem -t 64 -x ont2d -Y -E1 to produce *Figure 2a*.

DNA from Proband 13 was similarly extracted and sequenced by UCL LRS facility. Alignment was initially obtained with dorado v0.9.1.

For all samples, the region encompassing *SORD* and *SORD2P* underwent a further re-alignment with VACmap and processed with Sniffles2 to confirm the presence of the inversion. The VACmap alignment was also used to produce *Figure S5*.

### Optical Genome Mapping (OGM)

Ultra-high molecular weight DNA was extracted from blood using the ‘Blood and cell culture DNA isolation kit’ according to the manufacturer’s protocol. Following Qubit Fluorometer (Invitrogen) quantification, 750 ng DNA per sample was labelled using Bionano Prep DLS Labelling Kit. All samples were loaded onto Saphyr chips for linearisation and imaging and processed on a Bionano Saphyr machine. We ran the Bionano *de novo* assembly and used variant annotation pipelines to generate whole-genome assembly contigs, which span whole-chromosome arms and detect variants larger than 500 bp. Consensus contigs and SVs were manually visualised with Access v.3.7. All samples were run at nominal coverage of at least 100×.

### Hi-C and Fit-Hic analysis

The Hi-C experiment was performed on fibroblasts from Proband 1 carrying a *SORD*/*SORD2P* inversion following the protocol of Dimartino et al., 2024 [33]. In brief, fibroblasts were cultured and grown to confluence to obtain approximately one million cells for fixation in 2% formaldehyde. Fixed cells were then lysed and digested using DpnII enzyme (R0543S, New England BioLabs, Ipswich, MA, USA). After digestion, restricted fragments were marked incorporating biotin-14-dATP during overhang fill-in (19524-016, Thermo Fisher Scientific, Waltham, MA, USA) and proximal ends were ligated overnight using T4 DNA ligase (M0202, New England BioLabs). To remove proteins holding interacting loci in close proximity, we reversed formaldehyde cross-linking with proteinase K digestion in the presence of 10% SDS followed by a 4 h incubation at 68°C in 0.5M NaCl. DNA was purified by sodium acetate and ethanol precipitation. To prepare NGS libraries, biotinylated DNA was sonicated using Bioruptor Pico sonicator (Diagenode, Liège, Belgium), pulled down using Dynabeads MyOne Streptavidin T1 beads (65602, Thermo Fisher Scientific), end-repaired and phosphorylated using T4 DNA polymerase and T4 PNK (M0203 and M0201L, New England BioLabs). Sequencing adaptors and indexes were added to DNA fragments using NEBNext Multiplex Oligos for Illumina kit (E7335 and E7500, New England BioLabs) and NEBNext Ultra II Q5 Master Mix (M0544, New England BioLabs). Double size selection of PCR products was performed using Agencourt AMPure XP beads (A63881, Beckman Coulter, Brea, USA). Three libraries per case (technical replicates) were sequenced for ∼240 million fragments each in a 100 bp paired-end run on a NextSeq 2000 (Illumina, San Diego, CA, USA).

Hi-C data were processed using HiC-Pro v.3.1.0 with default options to obtain the coordinates of all valid interactions, starting from the raw fastq files. Reads were mapped to the GRCh37/hg19 reference build. HiC-Pro outputs from the three technical replicates were merged and converted to Juicer format using an HiC-Pro accessory script. We used Juicer Tools v.1.22.01 and Straw v.0.0.8 to normalize Hi-C matrices with the Knight-Ruiz (KR) matrix balancing algorithm and extract chromosome 15 cis interactions at 5 kb resolution, respectively. The Hi-C map was visualized as heatmaps rotated by 45°. Statistically significant contacts have been highlighted using Fit-Hi-C v.2.0.8 [31] at 5kb resolution and FDR threshold of 0,05. FitHiC outputs were visualized on UCSC Genome Browser.

### Breakpoint-Specific Polymerase Chain Reaction (PCR)

We designed two forward primers matching regions proximal, albeit external, to the inversion breakpoint (F2: 5’-TCACGCCATTCTCTCACCTCAG-3’; F: 5’-GGCAGGAGAATGGCTTGAACAC-3’). Considering the high sequence homology of the region, we designed primers that unambiguously align to either *SORD* (F primer) or *SORD2P* (F2 primer) by leveraging sequence variation within divergent Alu elements. The two primers are normally 226 kb apart and oriented in the same forward direction, so that they are unable to generate any PCR product. In the presence of the inversion, the F and F2 primer can align in opposite orientation and generate a PCR product of approximately 3.2 Kb. Long-range PCR amplification reactions were performed by using Veriti 96 Well Thermal Cycler, Applied Biosytems. The mix composition included DNA 1 µL (100 ng/µL), F primer 0,5 µL (10 µM), F2 primer 0,5 µL (10 µM), 2X Phusion Master Mix with HF Buffer 5 µL, Betaine 5 M 2 µL, H_2_O 1 µL. PCR conditions were as follows: 30 s at 98°C (initial denaturation) followed by 35 cycles of 10 s at 98°C; 30 s at 60°C; 3 min at 72°C, with a final extension for 5 min at 72°C. After long-range PCR amplification, agarose electrophoresis was performed. The breakpoint-specific PCR was employed to screen for the *SORD*/*SORD2P* inversion in genomic DNA isolated from blood samples.

### Demographic, clinical, and neurophysiological data of CMT patients with the *SORD*/*SORD2P* inversion

Demographic, clinical, and neurophysiological characteristics of CMT patients with the *SORD*/*SORD2P* inversion were collected. Disease severity was classified using the validated CMT Examination Score version 2 (CMTESv2), which categorizes severity as mild (CMTES 0 to 7), moderate (CMTES 8 to 16), or severe (CMTES 17 to 28) [34].

### Analysis of chromatin interactions of gene/pseudogene pairs

We investigated the genomic architecture of the *SORD* and *SORD2P* loci, as well as the other gene/pseudogene inversions, by analyzing in situ Hi-C and Micro-C XL tracks from the UCSC Genome Browser [29-30]. These tracks display heatmaps of chromatin interactions between DNA regions from *in situ* Hi-C and Micro-C XL experiments on the H1 embryonic stem cells and HFFc6 (foreskin fibroblasts) cell lines. Thus, high scores are represented by more intense colors and correspond to closer proximity. Both techniques provide a genome-wide and detailed picture of chromatin structure, by a cross-linking of the contact points, followed by digestion with MNase (Micro-C) or specific restriction enzymes (Hi-C), and mapping both ends of the resulting reads to a reference genome. Due to the increased density of MNase cutting sites, Micro-C reaches higher resolutions than Hi-C (<1 kbp vs 5 kbp). The heatmap visualization is obtained by binning and counting the number of mapped split-reads in each bin. It is important to note that any sequence with ambiguous mapping is excluded from the analysis. This is particularly relevant in our context, as it rules out the possibility of false signals due to perfect homology.

Statistically significant interaction counts were obtained with FitHic2, on the HFFc6 cell line data available at the 4dnucleome data portal [35]. All analyses were run at the maximum available resolution (1000 bp). Interaction data are provided by 4dnucleome in .hic format, which needs to be preprocessed with scripts (included in FitHic2 package) to generate intermediate complementary files (a contact file, a fragment file and a bias file, using createFitHiCContacts-hic.py, createFitHiCFragments-fixedsize.py and HiCKRy.py, respectively). The total counts for candidate pairs and control pairs were then obtained using a custom python script, including only fragment pairs found to be in statistically significant chromatin contact after Bonferroni correction (p < 1e-6).

## Data Availability

All data produced in the present study are available upon reasonable request to the authors.

## Supplementary material

### Supplementary Tables

Supplementary Table S1: Complete list of gene–pseudogene pairs arranged in inversion-permissive orientation across the human genome. The table includes the names of the identified pseudogene and their corresponding parent gene, together with the chromosomal coordinates and strand orientation of both loci.

Supplementary Table S2: List of parent genes from inversion-permissive gene–pseudogene pairs with known disease associations. This table lists genes from S1 that are linked to human diseases according to the OMIM database, along with information about associated phenotype and mode of inheritance.

Supplementary Table S3: Initial set of candidate inversions detected from long-read sequencing data in the 1000 Genomes Project through our custom pipeline and filtering criteria. Each row represents a distinct inversion event detected in a single sample. For each event, the parent gene, pseudogene, 1000 Genomes sample ID, genomic coordinates of the two breakpoints, and the number of supporting split-reads are provided. This dataset constitutes the filtered call set prior to our manual curation.

Supplementary Table S4: Demographic, clinical, and neurophysiological characteristics of patients with CMT/dHMN carrying the SORD/SORD2P inversion. The table reports clinical information for affected individuals identified as carriers of the SORD/SORD2P inversion. Abbreviations: dHMN, distal hereditary motor neuropathy; CMT, Charcot–Marie–Tooth disease; DTRs, deep tendon reflexes; AFOs, ankle foot orthoses; CMAP, compound muscle action potential; MCV, motor conduction velocity; SNAP, sensory nerve action potential; SCV, sensory conduction velocity; MRI, magnetic resonance imaging; N, normal; –, absent; +, present; ↑, increased; ↓, reduced; NA, not available.

### Supplementary Figures

Supplementary Figure 1: Long-read sequencing (LRS) reveals a distinct pathogenic inversion involving SORD and SORD2P in a CMT patient negative to PCR screening for the common inversion allele. i) Integrative Genomics Viewer (IGV) snapshot showing the split-reads marking the inversion breakpoints localization between exon 3 and exon 4 of SORD and exon 4 and exon 5 of SORD2P. ii) Schematic comparison illustrating the relative positions of the patient’s inversion versus the recurrent pathogenic inversion.

## Acknowledgments/Funding

The work is supported by Charcot-Marie-Tooth Association (SR-202504 to AC), AFM-Telethon (28813 to AC), European Reseach Council Starting grant (101165557 to AC), National Ataxia Foundation,Medical Research Council (MR/T001712/1 to AC), Fondazione Cariplo (grant n. 2019-1836 to AC), Fondazione Regionale per la Ricerca Biomedica (1751723 to AC), Muscular Dystrophy UK (24GRO-PG24-0719-1 to JS and AC). SZ is supported by NIH (5R01NS105755) and Muscular Dystrophy Association. AB thanks the European Academy of Neurology (EAN) for supporting him with a Research fellowship 2025. GA thanks the Peripheral Nerve Society for supporting him with a Clinical Training Fellowship 2025. SM thanks Ricerca Corrente RCR-2025 to Fondazione IRCCS Istituto Neurologico Carlo Besta for grant support. JS is supported by Medical Research Council (MR/Y010949/1).

