## Supplementary figures and images for "Gene-Pseudogene Inversions as a Hidden Source of Missing Heritability"

### Supplementary Figure 1

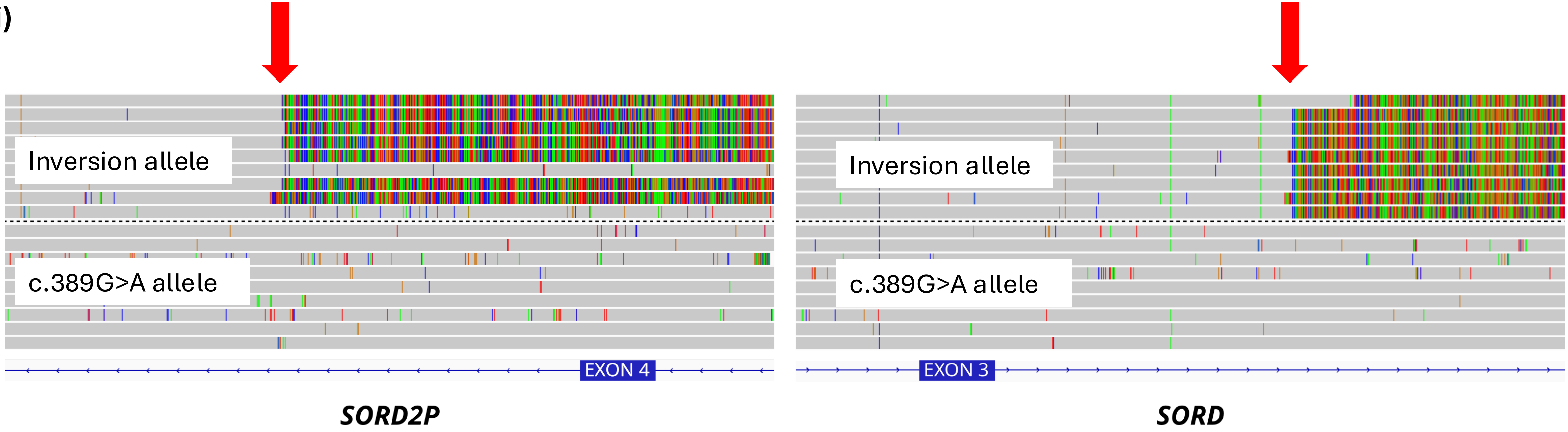

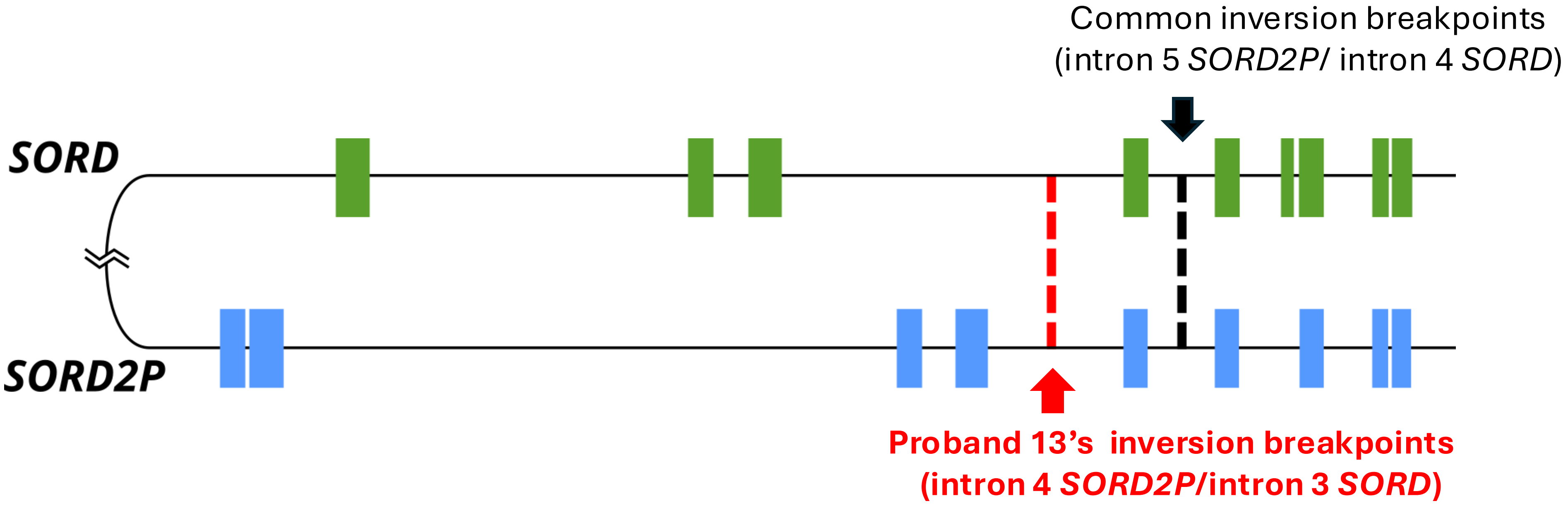
